# Unilateral cathodal transcranial direct current stimulation over the parietal area modulates on postural control depending with eyes open and closed

**DOI:** 10.1101/2022.05.16.22275178

**Authors:** Shinichiro Oka, Takuro Ikeda, Tsubasa Mitsutake, Katsuya Ogata, Yoshinobu Goto

## Abstract

Cathodal transcranial direct current stimulation (C-tDCS) is generally assumed to inhibit cortical excitability. The parietal cortex contributes to multisensory information processing in the postural control system, and this processing is proposed to be different between the right and left hemispheres and sensory modality. However, previous studies did not clarify whether the effects of unilateral C-tDCS of the parietal cortex on the postural control system differ depending on the hemisphere. We investigated the changes in static postural stability after unilateral C-tDCS of the parietal cortex. Ten healthy right-handed participants were recruited for right- and left-hemisphere tDCS and sham stimulation, respectively. The cathodal electrode was placed on either the right or left parietal area, whereas the anodal electrode was placed on the contralateral forehead. We evaluated static standing balance by measuring the sway path length, mediolateral (ML) sway, anteroposterior (AP) sway, sway area, and the sway path length per unit area (L/A) after 15-minute C-tDCS under eyes open (EO) and closed (EC) conditions. C-tDCS over the right hemisphere significantly increased the sway path length, ML sway, and sway area in the EO condition. In contrast, C-tDCS over the left hemisphere significantly increased the L/A in both the EC and EO condition. These results suggest that the right parietal region contributes to static standing balance through chiefly visual information processing during the EO condition. On the other hand, L/A increase during EC and EO by tDCS over the left parietal region depends more on somatosensory information to maintain static standing balance during the EC condition.

## Introduction

Postural stability depends on the integration of multisensory inputs, such as vision, vestibular, and somatosensory perception, to produce motor output (1). These sensory systems are integrated by the vestibular nuclei and parietal association area of the cerebral cortex to induce postural reflexes and voluntary movements to adapt to the external environment (1).

Brain imaging studies have shown that the parietal lobe is activated by various stimuli, including visual, vestibular, and somatosensory stimuli. The posterior parietal cortex (PPC) has been reported to be involved in information processing in the brain in relation to the integration of these multisensory systems (2). In previous studies, the right parietal area was reported to be activated by visual inputs such as optokinetic stimulation and fixation of visual targets (3) (4), while the left PPC was activated by somatosensory inputs such as light touch from a stable external spatial reference (5), vestibular inputs such as caloric stimulation (6), or galvanic vestibular stimulation (GVS) (7). Sensory processing and integration in the PPC have been shown to be dominated by the right hemisphere (3, 8-10).

In clinical studies, strokes that affect one or more postural control networks (visual, vestibular, and somatosensory) are known to present with lateropulsion (pusher syndrome) (11). Lateropulsion is characterized by a contralesional bias in posture with stroke, active resistance to postural correction to upright vertical (12), and weight-bearing asymmetry (WBA) (13). In particular, patients with lateropulsion and right parietal lesions show delayed functional recovery, necessitating prolonged rehabilitation efforts (14). This is attributable to the fact that WBA in lateropulsion patients is related to many factors, including motor deficits, sensory deficits, and spatial neglect (15). Therefore, clinical studies investigating the relationship between parietal lobe dysfunction and standing postural control in patients with lateropulsion are limited.

Transcranial electrical stimulation (tDCS) has been recently used to investigate the pathogenesis of brain dysfunction and develop neurorehabilitation programs. tDCS, a non-invasive electrical stimulation method that induces excitatory changes in the corticospinal circuitry, can be used to modulate cortical excitability by applying a weak current to an electrode attached to the head (16). In a previous study, bilateral tDCS to the parietal area modulated postural adaptation after tilting, suggesting that brain information processing in the parietal cortex contributes to standing posture control (17). However, there was no difference in the effects on postural control depending on the polarity of stimulation by bilateral tDCS in the parietal region in that study (17). That results might be due to bilateral stimulation. Therefore, transient functional inhibition by unilateral tDCS may clarify the relationship between left and right parietal functions and standing posture control. In addition, the influence of the sensory system on standing posture control has been investigated by varying visual conditions(18). Therefore, the differences in the effect of brain information processing in the left and right parietal cortices on standing posture control could be compared between eyes open (EO) or closed (EC) conditions. Clarification of the functional relationship between brain dominance and standing posture control can reveal the influence of brain dysfunction on WBA in lateropulsion patients and lead to the development of neurorehabilitation protocols for parietal lobe dysfunction.

In previous studies, unilateral cathodal (C)-tDCS has been used to modify information processing in the hemisphere (9) (19). On the other hand, bilateral tDCS has been used in studies of bilateral cerebral hemispheric effects (9) (20). Therefore, we aimed to induce transient functional suppression of the unilateral parietal cortex by C-tDCS and investigate the relationship between sensory information processing in the brain and postural control under the EO and EC conditions to differentiate the dependence of visual information.

## Methods

### Participants

A total of ten right-handed healthy young adults (5 females; mean ± SD, 21.4 ± 0.8 years old) participated in this study. None of the participants had a history of neurological, orthopedic, or other medical problems. All participants gave written informed consent in accordance with the Declaration of Helsinki. The study was approved by the Ethics Committee of International University of Health and Welfare (15-Ifh-18).

### tDCS

The participants sat on a comfortable chair in a quiet room during stimulation. C-tDCS was delivered using a battery-driven current stimulator (DC Stimulator-Plus; NeuroConn GmbH, Ilmenau, Germany) through two rubber electrodes with sponge pads soaked in saline solution and affixed using a Velcro support. C-tDCS was applied at 1.5 mA for 15 minutes (the impedance was maintained below 5 kΩ), with 5 s of ramping up and down, in accordance with the protocol described in previous studies (9) (17). The positions of the stimulation electrodes were adopted from previous studies (9, 19, 20). The tDCS cathodal electrode (surface area: 35 cm^2^, 7 × 5 cm) was placed at P3 or P4 according to the International 10-20 system, and the anodal electrode (surface area: 35 cm^2^, 7 × 5 cm) was placed in the contralateral orbit. These electrode positions were selected to affect the parietal cortex (9). For sham stimulation, tDCS was applied for 30 s at the beginning of the 15-min period. After stimulation, all the participants were asked to report whether they experienced any tDCS-induced sensations.

### Postural control task

Postural control was assessed under bipedal static stance conditions by using a stabilometer (Twingravicoder G-6100; Anima Co. Ltd., Chofu, Japan). The system recorded the center of foot pressure (COP) trajectories over time, in both the mediolateral (COP-X) and anteroposterior (COP-Y) directions, at a sampling frequency of 20 Hz for one minute. The measurements were performed under EO or EC conditions. The participants stood without shoes or feet together. Each participant was instructed to stand as still as possible while looking forward and keeping the arms relaxed at the sides. In the EO condition, participant was instructed to fixate on the fixation point with a with a diameter of about 2 cm was placed 2 m in front of the them at the eye level.

### Experimental procedures

In this randomized, single-blind study, sessions with different C-tDCS conditions were separated by at least two days. The experimental procedure is shown in Fig. 1. C-tDCS intervention was tested under three conditions: sham, right:P4 cathodal, and left:P3 cathodal. The participants were seated on a chair, and tDCS electrodes were placed on the parietal area and contralateral orbit. They subsequently underwent COP measurement without stimulation (baseline) followed by COP measurements in each intervention. During intervention the subjects involved sitting on the chair for about 20 minutes. The order of interventions was randomized, but the COP measurements were performed in the EO condition first, followed by the EC condition.

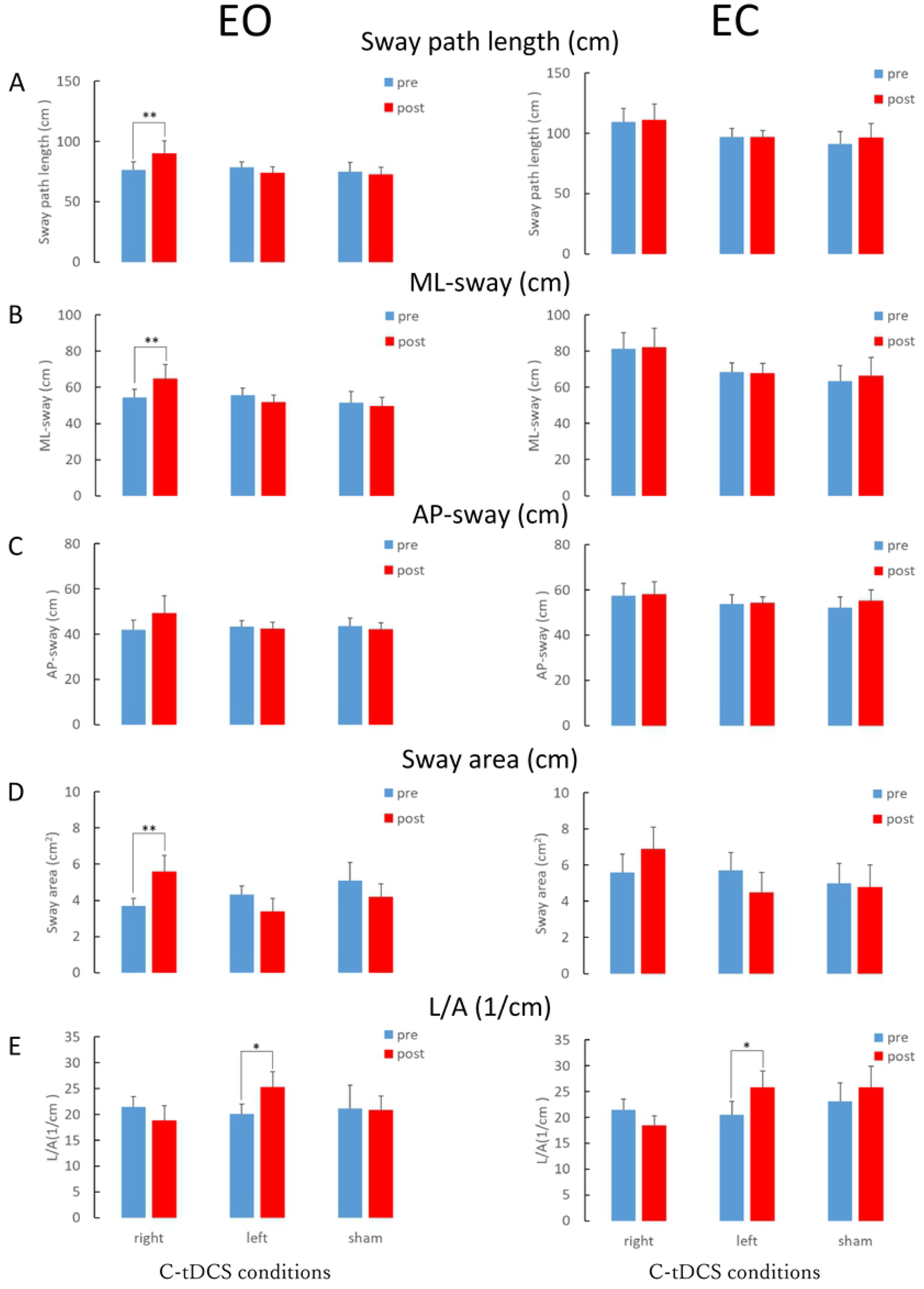

### Analysis of the postural control task

The locus in the COP was converted to values of statistical indices such as sway path length, mediolateral (ML) sway, anteroposterior (AP) sway, sway area, and sway path length per unit area (L/A). The ML sway integrated the movement of the COP in the left-right direction. The AP sway integrated the amount of COP movement in the front-back direction. L/A is considered a parameter for fine control of standing posture by proprioceptive reflexes (21) (22).

### Statistical analyses

The sway path length, ML sway, AP sway, sway area, and L/A were calculated at baseline (Pre) and after stimulation (Post) in each condition. To evaluate the effects of C-tDCS on pre- and post-offline trials, each parameter was compared using two-way repeated-measures analysis of variance (ANOVA) with factors of intervention (right, left, and sham) and time (pre, post). Preliminary testing for normality with the Shapiro–Wilk test showed that the data were normally distributed in all assessments. Sphericity was assessed using Mauchly’s test. When a significant difference was observed in the interaction in repeated-measures ANOVA, a post-hoc evaluation was performed using a paired t-test. Comparisons of baseline values among the experimental conditions were analyzed using one-way repeated-measures ANOVA. The data are presented as the mean ± standard error of the mean. Moreover, the effect sizes were evaluated according to standardized size-effect indices of partial eta-squared (η_P_^2^) and Cohen’s d. Statistical analysis was performed using SPSS statistics (version 25.0 for Windows, IBM, Armonk, NY, USA). Statistical significance was set at p < 0.05. The EO and EC data were analyzed separately.

## Results

The results of the two-way repeated-measures ANOVA for EO are shown in Table 1 and Figure 1. No significant differences were observed in baseline sway path length, ML sway, AP sway, sway area, or L/A among the experimental sessions (sway path length, p = 0.925; ML sway, p = 0.831; AP sway, p = 0.930; sway area, p = 0.431; L/A, p = 938). A significant interaction was observed between intervention and time for sway path length (F (2, 27) = 4.740, p = 0.017, η_P_^2^ = 0.260). ML sway (F (2, 27) = 4.926, p = 0.015, η_P_^2^ = 0.267), and sway area (F (2, 27) = 9.624, p = 0.001, η_P_^2^ = 0.416). Post-hoc comparisons with paired t-tests revealed that sway path length, ML sway, and sway area increased significantly only after right hemisphere stimulation (sway path length, p < 0.01, d = 0.51; ML sway, p < 0.01, d = 0.52; sway area, p < 0.05, d = 0.83). L/A increased significantly after left-hemisphere stimulation (p < 0.05, d = 0.67), but no differences were observed in the sham stimulation (Fig. 1).

**Table1.** Effect of C-tDCS in sway path length, ML-sway, AP-sway, and L/A with EO condition

| Average±SE | Pre(right) | Post(right) | d | Pre(left) | Post(left) | d | Pre(sham) | Post(sham) | d |
| --- | --- | --- | --- | --- | --- | --- | --- | --- | --- |
| Sway path length (cm) | 76.3±6.7 | 90.3±10.3** | 0.51 | 78.5±4.5 | 74.2±4.7 | 0.29 | 75.0±7.4 | 72.6±5.9 | 0.12 |
| ML sway (cm) | 54.5±4.5 | 65.0±7.8** | 0.52 | 55.7±3.7 | 51.7±3.9 | 0.33 | 51.6±6.0 | 49.6±4.9 | 0.12 |
| AP sway (cm) | 41.8±4.3 | 49.2±5.5 | 0.48 | 43.4±2.5 | 42.3±2.9 | 0.13 | 43.5±3.6 | 42.1±2.8 | 0.14 |
| Sway area (cm<sup>2</sup>) | 3.7±0.4 | 5.6±0.9** | 0.83 | 4.3±0.5 | 3.4±0.7 | 0.54 | 5.1±1.0 | 4.2±0.7 | 0.32 |
| L/A (1/cm) | 21.5±1.9 | 18.9±2.8 | 0.34 | 20.1±1.9 | 25.0±2.9* | 0.67 | 21.2±4.4 | 20.8±2.8 | 0.03 |
| Two-way repeated measures ANOVA | Time |  |  | Intervention |  |  | Time × Intervention |  |  |
| | F(df) | $\eta_p^2$ | p | F(df) | $\eta_p^2$ | p | F(df) | $\eta_p^2$ | p |
| Sway path length | 0.829(1,27) | 0.030 | 0.371 | 0.579(2,27) | 0.041 | 0.567 | 4.740(2,27) | 0.260 | 0.017 |
| ML sway | 0.528(1,27) | 0.019 | 0.474 | 0.854(2,27) | 0.059 | 0.437 | 4.926(2,27) | 0.267 | 0.015 |
| AP sway | 0.817(1,27) | 0.029 | 0.347 | 0.210(2,27) | 0.015 | 0.812 | 2.550(2,27) | 0.159 | 0.097 |
| Sway area | 0.021(1,27) | 0.001 | 0.886 | 0.474(2,27) | 0.034 | 0.628 | 9.624(2,27) | 0.416 | 0.001 |
| L/A | 0.243(1,27) | 0.009 | 0.626 | 2.853(2,27) | 0.015 | 0.821 | 2.853(2,27) | 0.174 | 0.075 |

Multiple comparison (Intervention): Bonferroni, ns, simple main effect: \*\* $p < 0.01$ , \* $p < 0.05$
Effect size : $\eta_p^2 = 0.010$ ~small effect, $\eta_p^2 = 0.060$ ~medium effect, and $\eta_p^2 = 0.140$ ~large effect
$d = 0.20$ ~small effect, $d = 0.50$ ~medium effect, and $d = 0.80$ ~large effect
ANOVA: analysis of variance, AP: antio-posterior, ML: medio-lateral, L/A: sway path length per unit area

The results of the two-way repeated-measures ANOVA for EC are shown in Table 2. No significant differences were observed in baseline sway path length, ML sway, AP sway, sway area, and L/A between the experimental sessions (sway path length, p = 0.390; ML sway, p = 0.246; AP sway, p = 0.743; sway area, p = 0.853; L/A, p = 807). A significant interaction was observed between the intervention and time for L/A (F (2, 27) = 3.429, p = 0.047, η_P_^2^ = 0.203). Post-hoc comparisons with paired t-tests revealed that L/A increased significantly (p < 0.05, d = 0.57) after left-hemisphere stimulation, but not after the right hemisphere or sham stimulation (Fig. 1).

**Table2.** Effect of C-tDCS in sway path length, ML-sway, AP-sway, and L/A with EC condition

| Average±SE | Pre(right) | Post(right) | d | Pre(left) | Post(left) | d | Pre(sham) | Post(sham) | d |
| --- | --- | --- | --- | --- | --- | --- | --- | --- | --- |
| Sway path length (cm) | 109.6±11.1 | 111.4±12.6 | 0.05 | 96.9±7.1 | 97.0±5.4 | 0.02 | 90.9±10.4 | 96.2±11.9 | 0.15 |
| ML sway (cm) | 81.4±8.7 | 82.3±10.3 | 0.03 | 68.5±5.1 | 67.8±5.3 | 0.04 | 63.3±8.6 | 66.4±10.2 | 0.11 |
| AP sway (cm) | 57.3±5.5 | 58.1±5.6 | 0.05 | 53.7±4.1 | 54.4±2.5 | 0.07 | 52.1±4.8 | 55.2±4.9 | 0.21 |
| Sway area (cm<sup>2</sup>) | 5.6±1.0 | 6.9±1.2 | 0.35 | 5.7±1.0 | 4.5±1.1 | 0.45 | 5.0±1.1 | 4.8±1.2 | 0.04 |
| L/A (1/cm) | 21.6±2.0 | 18.4±1.9 | 0.53 | 20.5±2.6 | 25.8±3.2* | 0.57 | 23.1±3.6 | 25.8±4.1 | 0.22 |
| Two-way repeated measures ANOVA | Time |  |  | Intervention |  |  | Time × Intervention |  |  |
| | F(df) | $\eta_p^2$ | p | F(df) | $\eta_p^2$ | p | F(df) | $\eta_p^2$ | p |
| Sway path length | 1.184(1,27) | 0.042 | 0.286 | 0.825(2,27) | 0.058 | 0.449 | 0.704(2,27) | 0.050 | 0.503 |
| ML sway | 0.467(1,27) | 0.017 | 0.500 | 1.208(2,27) | 0.082 | 0.314 | 0.462(2,27) | 0.033 | 0.635 |
| AP sway | 1.576(1,27) | 0.055 | 0.220 | 0.234(2,27) | 0.017 | 0.793 | 0.409(2,27) | 0.029 | 0.668 |
| Sway area | 0.015(1,27) | 0.001 | 0.903 | 0.557(2,27) | 0.040 | 0.579 | 2.959(2,27) | 0.180 | 0.069 |
| L/A | 1.350(1,27) | 0.048 | 0.255 | 0.700(2,27) | 0.049 | 0.505 | 3.429(2,27) | 0.203 | 0.047 |

Multiple comparison (Intervention): Bonferroni, ns, simple main effect: \*\* $p < 0.01$ , \* $p < 0.05$
Effect size : $\eta_p^2 = 0.010$ ~small effect, $\eta_p^2 = 0.060$ ~medium effect, and $\eta_p^2 = 0.140$ ~large effect
d = 0.20 ~small effect, d = 0.50 ~medium effect, and d = 0.80 ~large effect
ANOVA: analysis of variance, AP: antero-posterior, ML: medio-lateral, L/A: sway path length per unit area

## Discussion

In the present study, C-tDCS on the right parietal area increased sway path length, ML sway, and sway area during the EO condition. In contrast, C-tDCS to the left parietal area increased the L/A during the EO and EC conditions. A previous study reported that bilateral tDCS to the parietal area modulates postural adaptation after tilting (Young 2020), indicating that information processing in the parietal cortex contributes to control of the standing posture. The modulation of standing posture control by unilateral C-tDCS to the parietal area in the present study supports the findings of a previous study and extends our knowledge by revealing the differential effect of C-tDCS depending on the stimulus side of the parietal area and the visual condition (EO or EC). Therefore, the current study indicates a hemispheric difference in the effects of the parietal lobe on postural control through the integration of multisensory information.

### C-tDCS on the right parietal area impaired postural control during EO

C-tDCS on the right parietal area increased the sway path length, ML sway, and sway area during the EO condition but not during EC. Therefore, C-tDCS over right parietal area is assumed to impair postural control in a state of higher dependence on visual information processing. Static EO standing is controlled by inputs from the visual, somatosensory, and vestibular senses. However, cortical activity during visual and vestibular input has been shown to have a reciprocal inhibitory effect (Naito et al., 2003, Dietrich et al., 2003). Therefore, EO static standing balance is considered to be controlled by the visual and somatosensory systems, as vestibular information processing in the brain is suppressed. The visuospatial information related to standing posture control during EO is processed in the PPC, with a predominance on the right hemisphere (10) (2). A previous fMRI study reported that vertical/horizontal lines increased neural activity in the superior and inferior parietal cortices bilaterally, although the increase was observed predominantly on the right (23).

Bilateral t-DCS over the parietal areas (left-anodal, right-cathodal) induced visual mislocalization to the right (20). TMS of the right PPC has been reported to inhibit the coding of positional information obtained by gazing to visual stability (24). We speculate that C-tDCS to the right parietal may have increased ML-sway by suppressing the processing of the vertical line from the floor to the fixed viewpoint at eye level. In addition, stroke patients with right PPC lesions have been reported to show general spatial memory impairments (25). Visuospatial information requires a dynamic spatial map that integrates information sampled from retinal images, and is maintained and updated for each new gaze position (remapping process) (10). Above all, these studies suggest that C-tDCS to the right parietal area were suppressed right parietal cortex and modulated the vertical line perception by the fixation point leading visual instability to update spatial information during standing, which resulted in the sway path length and ML-sway increase.

The sway area reflects not only visual but also proprioceptive function during postural control, and in the EO condition, the contribution of the sway area has been shown to be higher for proprioceptive function than visual field testing scores (18). An fMRI study on foot positional perception suggested that the significant regions responsible for position sense are in the right parietal and frontal cortices (26). In a combined visual and proprioceptive sensory stimulation task experiment, Christensen et al. reported that visually guided self-generated ankle movements activated the PPC (27). Convento et al. revealed that anodal tDCS to the right temporoparietal junction (TPJ) modulates proprioceptive sensory alignment and illusory perception, suggesting that the right TPJ and related areas contribute to the integration of vision and touch (8). Therefore, the increase in sway area with EO after C-tDCS to the right parietal area in the present study may be also due to modulation of the integration of visual and proprioceptive information.

### C-tDCS on the left parietal area impaired postural control during EC and EO

We found that C-tDCS on the left parietal area showed a significant interaction (time × intervention) and large effects on L/A in the EC condition. The L/A ratio in the EO and EC conditions increased after C-tDCS on the left parietal area, which suggested that postural instability by C-tDCS on the left parietal area is not dependent on visual information processing. Sway area in standing balance with EO and EC in healthy older adults was reported to be contributed by somatosensory rather than age(18), indicating that somatosensory perception plays an important role in standing postural control at firm surface with or without vision. Furthermore, L/A, sway length divided by sway area, is considered a parameter reflecting the fine control of standing posture by proprioceptive reflexes (21) (22). L/A is used as an indicator of somatosensory-derived fine body sway(28) (29). In clinical studies, L/A has been used to assess body sway in postoperative patients with cervical myelopathy (28), and preventing the potential risk of falls and body sway after taking antidepressants (29). In addition, the frequency band of body sway has been shown to be related to sensory information processing for standing posture control. The frequency of body sway during static standing has been reported to have an average frequency of 0.11 ± 0.07 Hz during the EO condition (30), and the median frequency increases during the EC condition (31). In the standing balance task, a reflex response coherent with perturbation was seen in the soleus EMG at frequencies up to 5 Hz, with maximal coherence at 1.0-2.0 Hz (32), and highest for the 1- to 2-Hz stochastic vestibular stimulation signal (33). In particular, participants with higher L/A had a higher power spectrum at 2-5 Hz (21), which is considered a parameter of fine control of standing posture by proprioceptive reflexes (32). The static standing posture in the EC condition is controlled by somatosensory and vestibular information (34). However, postural instability due to vestibular dysfunction is assessed by the COP in the foam rubber (35). Therefore, L/A represents postural control in a proprioceptive manner and is likely to be less influenced by the vestibular function. Furthermore, previous studies on somatosensory information processing in the brain showed that the left PPC is activated during a crossed-hand posture (36) and light touch with EC (5). C-tDCS of the left PPC was also reported to increase the limb position drift away from the defined target without visual feedback (19). The left inferior parietal lobule was activated during both hand-object illusions with the right and left hands, and the activity was greater than that in the right corresponding parietal region, suggesting a dominant role for the left hemisphere (37). Therefore, the L/A increase after C-tDCS of the left parietal lobe may be attributed to suppression of somatosensory information processing, contributing to high frequency of posture control independent of in the EC and EO.

### Limitations

This study had several limitations. First, the effects of tDCS on vision, vestibular perception, and somatosensory perception in the left and right parietal regions have not been investigated. Further studies are needed to clarify the effects of tDCS on the left and right parietal areas on vision, vestibular perception, and somatosensory perception. Second, the sample size was relatively small. Variability between individuals in response to transcranial direct current stimulation (tDCS) is a commonly reported issue in tDCS literature in recent years (38). However, the sample size was within the average range reported in other studies (39) (40) (41). In addition, the effect sizes in the current study were medium to large, implying that the effects of unilateral C-tDCS on postural control were robust. The differences between the C-tDCS and sham conditions were also not significant for all items. Finally, we used rectangular stimulation electrodes (5 × 7 cm), which did not allow focal stimulation (42). Therefore, co-stimulation of the cortical areas adjacent to the PPL is difficult to rule out.

## Conclusions

This study investigated the effects of unilateral C-tDCS on the parietal area during postural control. C-tDCS on the right parietal area significantly increased sway length, ML sway, and sway area during the EO conditions, while that over the left hemisphere increased L/A during the EO and EC conditions. Thus, the right parietal area controls body sway using visual and proprioceptive information, whereas the left parietal area controls high-frequency body sway using proprioceptive information during the EC condition. In future studies, we hope to clarify the relationship between information processing in the brain of the parietal cortex and sensory systems and develop neurorehabilitation protocols to improve balance based on the function of the parietal cortex.

## Data Availability

All relevant data are within the manuscript and its Supporting Information files.

